# No effect of resistance exercise on antibody responses to influenza vaccination in older adults: a randomized control trial

**DOI:** 10.1101/2020.08.19.20178442

**Authors:** Mahmoud T. Elzayat, Melissa M. Markofski, Richard J. Simpson, Mitzi Laughlin, Emily C. LaVoy

## Abstract

Older adults are at elevated risk for morbidity and mortality caused by influenza. Vaccination is the primary means of prophylaxis, but protection is often compromised in older adults. As acute eccentric resistance exercise mobilizes immune cells into muscle, it may enhance vaccination response. PURPOSE: Compare antibody responses to influenza vaccination in older adults who performed resistance exercise prior to vaccination to those who did not exercise. METHODS: 29 resistance training-naive older adults (20 women, 73.9 ± 5.3 years) were randomized to 1 of 3 groups: vaccination in the same arm that exercised (Ex-S), vaccination in the opposite arm that exercised (Ex-OP), and seated rest (No-Ex). Exercise was unilateral and consisted of 10 sets of 5 eccentric repetitions at 80% of the pre-determined concentric one repetition maximum. Lateral raises were alternated with bicep curls. No-Ex sat quietly for 25 min. Following exercise or rest, all subjects received the 2018 quadrivalent influenza vaccine (Seqirus Afluria) in the non-dominant deltoid. Antibody titers against the four influenza vaccine strains were determined by hemagglutinin inhibition assays at baseline, 6-, and 24-weeks post-vaccination. Group differences in antibody titers by time were assessed by restricted maximum likelihood mixed models. Fold-changes in antibody titers 6- and 24-weeks from baseline were compared between groups by Kruskal-Wallis tests. RESULTS: No significant group x time effects were found for any strain. Groups did not differ in fold-increase of antibody titers 6- and 24-weeks post-vaccination. Although seroconversion rates remained low, only one subject (Ex-S) reported flu-like symptoms 18 weeks post-vaccination. CONCLUSION: Acute arm eccentric exercise did not influence antibody titers to the influenza vaccine delivered post-exercise in older adults. More strenuous exercise may be required for exercise to act as an adjuvant. http://ClinicalTrials.gov Identifier: NCT03736759 U.S. NIH Grant/Contract: R03AG052778

## INTRODUCTION

The aging immune system leaves older adults more vulnerable to infections, including with influenza virus. Influenza infection is associated with the death of thousands of people in the United States each year, approximately 90% of which occur among older adults (≥65yrs)[1–3]. The primary means of protection against influenza is through annual vaccination with the seasonal influenza vaccine. Unfortunately, protective responses are impaired in this population, which likely accounts for the increased prevalence of infection in older adults [4–6]. One common approach to assess protection is measurement of influenza-specific antibody titers. Older adults exhibit lower antibody responses to the influenza vaccine leading to a clinical vaccine efficacy of just 17–53%, compared to 70–90% efficacy in younger adults [4,5]. Furthermore, antibody titers to influenza may decline more rapidly in older adults, meaning the effectiveness of the influenza vaccine may be further reduced if influenza exposure occurs late in the season [6]. This reduction in vaccination response is present even in older adults without chronic disease [7,8].

Considering the importance of generating protective immune responses against influenza infection, the development of strategies to improve vaccine responses among older adults is vital. Deficits in the antibody response to vaccines arise from age-related declines in the function of B-cells and T-cells [9,10], and potentially, antigen-presenting cells including dendritic cells [11]. Different formulations of the influenza vaccine for older adults have been proposed to enhance the protective levels of immune responses, such as the use of different adjuvants, different modes of administration, and/or increased antigen dose in the vaccine [12–15]. Although these strategies are typically more immunogenic than conventional non-adjuvanted influenza vaccines, they are also associated with greater injection site symptoms (e.g. erythema, swelling, and pain) and general systemic symptoms (e.g. malaise) post-vaccination [12–15]. While the safety profile is clinically acceptable, increases in the potential discomfort of vaccination could reduce vaccine uptake. Further, increases in the antigen dose in certain vaccine formulations require increased manufacturing capabilities and thus may limit available vaccine doses. Thus, there is a need to develop a simple, cost-effective approach with minimal side-effects to enhance influenza vaccine responses in older adults.

One potential method to enhance vaccine responses in older adults may be through localized resistance exercise designed to induce mild transient muscle damage. Eccentric exercise, or the application of tension as a muscle lengthens, reliably causes mild muscle damage and a local inflammatory response consisting of increased blood flow, vascular permeability, and immune cell invasion into the targeted muscle [16]. The inflammatory response is especially noted in resistance-training naïve individuals [16,17]. In young adults, resistance exercise involving eccentric contractions of the deltoid and biceps bracchi muscles immediately prior to inoculation improves antibody titer responses to vaccination [18–20]. However, a similar strategy has yet to be implemented in older adults who are more likely to benefit from these adjuvant effects of exercise. Moreover, it has not been determined from these published studies if the exercise effects are due to a local or a systemic response, as only the exercised arm has been inoculated. Eccentric resistance exercise also causes change in systemic immune parameters, such as increases in circulating leukocytes, altered expression of migration and adhesion molecules on neutrophils and monocytes, and increases in pro-inflammatory cytokines [16]. Therefore, exercise could act as an adjuvant through generalized immune upregulation.

The aim of this study was to determine the impact of a single bout of unaccustomed eccentric arm/shoulder resistance exercises performed immediately prior to inoculation on antibody responses to the seasonal influenza vaccine in older adults. We also aimed to compare the effect of identical bouts of exercise prior to vaccination in the inoculated arm and the non-inoculated arm. We hypothesized that vaccination in the arm that performed the eccentric resistance exercise would increase antibody titers at 6 weeks and 24 weeks post-vaccination relative to no-exercise and vaccination in the arm that did not exercise. These experiments will allow us to determine if a single bout of eccentric resistance exercise can be used as a simple and inexpensive vaccine adjuvant, with low side-effects, in community dwelling older adults. These experiments will also help differentiate between the local and systemic effects of eccentric exercise on vaccine enhancement.

## 2. MATERIALS AND METHODS

### 2.1 Study Design and Participants

This parallel randomized controlled trial (http://ClinicalTrials.gov Identifier: NCT03736759) compared vaccine responses at three time points across three groups (Figure 1). Data were collected October 2018 through April 2019 in Houston, Texas. Men and women aged ≥65yrs were recruited from the metropolitan region of Houston, Texas. Participants were screened to ensure they were non-frail (screened using Fried’s criteria [21]), non-smokers (>10 yrs), were not institutionalized, and met the American College of Sports Medicine criteria for participation in exercise [22]. Participants were excluded if they reported: i) a history of immune disease or vaccine-related allergies, ii) having a physician-confirmed influenza infection in the prior year, iii) regular use of medications known to affect the immune system, iv) were bedridden in the prior 3 months, v) engagement in resistance arm exercises in the prior 6 months, vi) an impairment limiting exercise or prohibiting informed consent, and vii) already vaccinated with the 2018 seasonal influenza vaccine. This population was expected to manifest age-attenuated responses to vaccination but be able to safely complete the bout of eccentric exercise. All participants were naïve to resistance training of the upper body to preclude the repeated bout effect potentially minimizing inflammatory responses [17]. This study was approved by the Institutional Review Board at the University of Houston (STUDY00000542).

**Figure 1.**
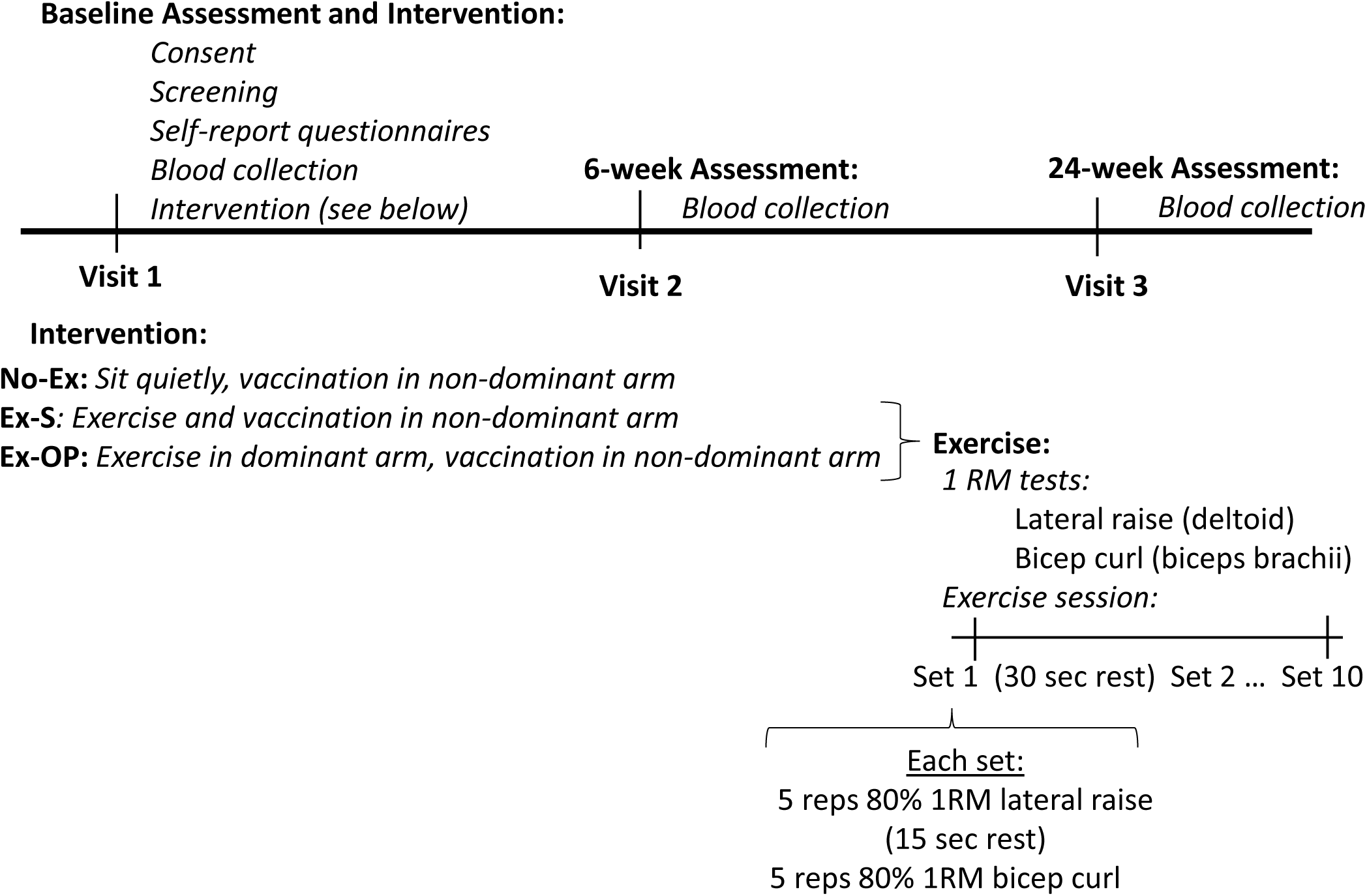
Experimental Design. No-Ex: Group that did not exercise, Ex-S: Group that received vaccine in the exercised arm; Ex-OP: Group that received vaccine in the opposite arm that was exercised; RM: repetition maximum; reps: repetitions.

Fifty-eight participants were initially screened for eligibility by phone or email; 16 did not meet inclusion criteria, and 12 declined to participate further or had schedule conflicts (Figure 2). Thirty participants were further screened in-person during the beginning of the first visit. Screening included a blood-pressure measurement, a survey of health behaviors and vaccine questions, the Mini-Mental State Examination, Fried’s Frailty Criteria (including handgrip strength determination using a hydraulic hand dynamometer), and the ACSM/AHA Exercise Readiness Questionnaire. One participant was excluded from further participation due to exclusion criteria. Twenty-nine participants provided informed consent and were randomly assigned to one of three groups at a 1:1:1 ratio, stratified by sex: (1) non-exercise inoculated control (No-Ex); (2) vaccination in the same arm that performed eccentric exercise of the deltoid and biceps bracchi (Ex-S); and (3) vaccination in the opposite arm that performed eccentric exercise of the deltoid and biceps bracchi (Ex-OP). Randomization was achieved via a random number string generated in Microsoft Excel following the recommendations for adaptive randomization by Hoare et al [23]. All participants completed Visit 1 in October 2018.

**Figure 2.**
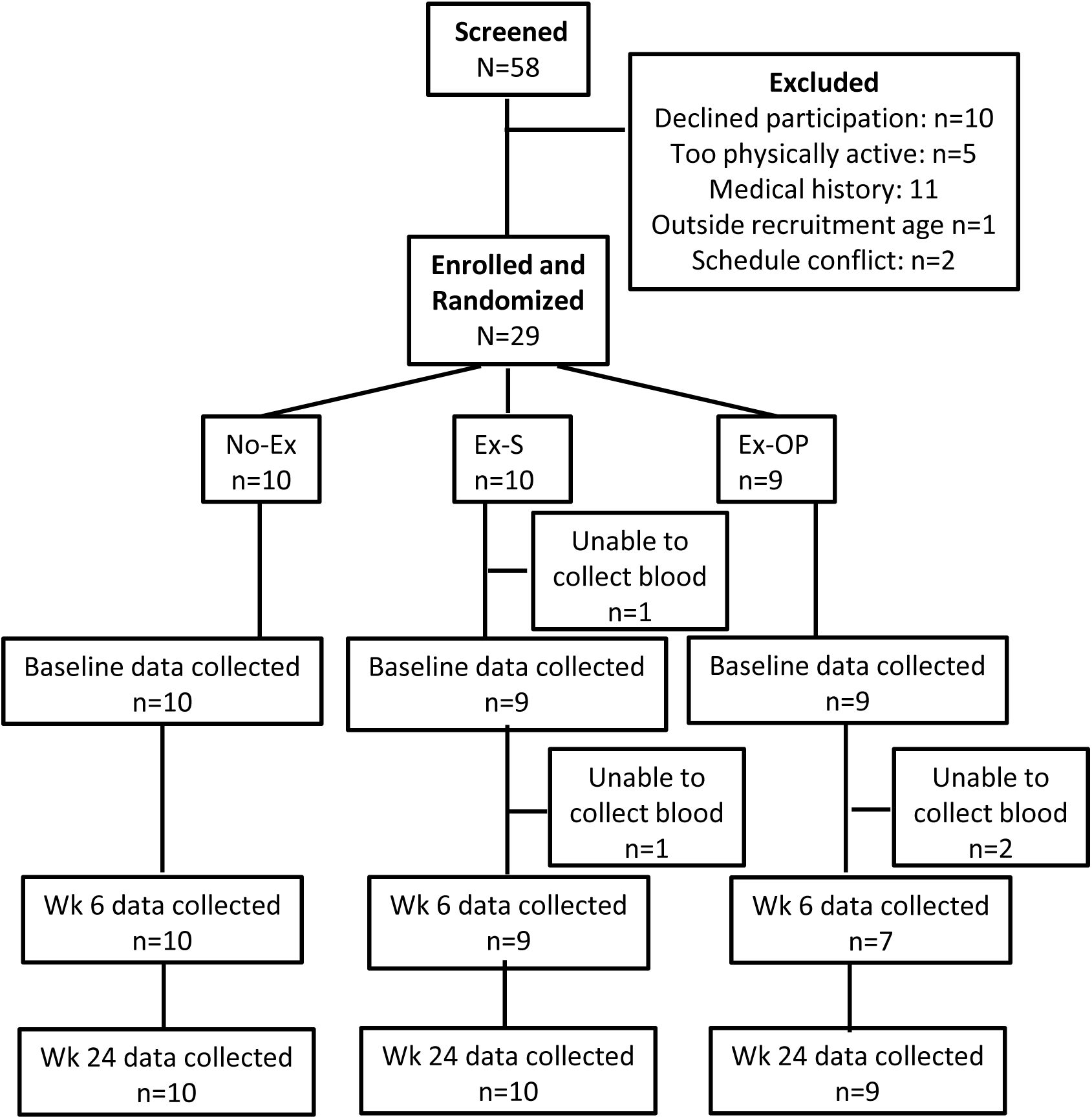
CONSORT flow diagram.

Participants completed two additional visits 6-weeks and 24-weeks post-vaccination to provide a blood sample. These visits were expected to correspond to peak antibody-responses (6-weeks post-vaccination) [24] and also indicate if protection was provided for the entire influenza season (24-weeks post-vaccination). Participants were compensated for their participation in each of the three visits with a gift-card.

### 2.2 Intervention and Vaccination

Following screening and consent process during Visit 1, participants completed questionnaires relating to life-stress and health behaviors (Perceived Stress Scale [25], General Health Questionnaire-28 [26], RAND 36-Item Health Survey [27], and a venous blood sample from the arm.

Participants assigned to Ex-S and Ex-OP completed single arm estimated 1 repetition maximum (1RM) tests for the bicep curl and lateral raise exercises. Briefly, participants completed 8–10 unassisted repetitions of the lateral raise exercise (shoulder abduction to 90° with elbow extension) and bicep curl exercise (elbow flexion) using dumb bells of increasing weight until 8 repetitions could not be completed with proper technique. The 1RM for each exercise was estimated using the Brzycki Formula [28].

After the experimental resistance exercise load was determined, participants completed 10 sets of 5 repetitions of the eccentric component of each movement at 80% of their estimated 1RM by lowering the dumbbell in a controlled manner over the course of 4 seconds. A study team member assisted with the concentric component of the exercise. Each experimental set consisted of one set of lateral raise exercise alternated with one set of bicep curl exercise. Weight was lowered if technique suffered to ensure that participants could complete the same number of repetitions. Participants rested 15 seconds after each set of lateral raise exercise (intra-set rest) and 30 seconds after each set of bicep curl exercise (inter-set rest). Participants in Ex-S completed the exercises in their non-dominant arm whereas Ex-OP completed the exercises in their dominant arm. Testing and exercise were completed in approximately 25 minutes. No-Ex participants rested quietly for 25 minutes.

Following exercise or seated rest, all participants were provided the quadrivalent influenza vaccine developed for the 2018–2019 influenza season (Afluria quadrivalent influenza vaccine; Seqirus). The quadrivalent vaccine included four virus strains: A/Michigan/45/2015 (H1N1), A/Singapore/INFIMH-16–0019/2016 (H3N2), B/Colorado/06/2017, and B/Phuket/3073/2013. Injections were given intramuscularly, directly into the deltoid of the non-dominant arm.

Participants were asked to report muscle soreness level using a visual analog scale before and after the injection, as well as each day for the next 7 days after injection via a scheduled phone call with a study team member (secondary outcome). Participants were contacted monthly for the remainder of the 6-month intervention to survey for flu-like symptoms (secondary outcome) and were asked to contact the study team if diagnosed with influenza.

### 2.3 Hemagglutinin Inhibition Assays

A primary outcome of this study was antibody titers to each influenza strain included in the vaccine. Venous blood was collected into 10 mL serum collection tubes (Vacutainer, BD) prior to vaccination (Baseline), 6 weeks-post vaccination, and 24 weeks-post vaccination. Serum was isolated by centrifugation from each sample within two hours of blood draw, and stored at –80°C. Following the final blood collection, all samples were shipped overnight on dry ice to a commercial laboratory (Southern Research, Birmingham AL) for the measurement of anti-influenza antibodies against the four influenza virus strains present in the vaccine. A standard microtiter hemagglutination inhibition assay was performed by researchers blinded to the intervention. Samples were analyzed in duplicate with a repeat performed as per standard operating procedure. Correctness of data was verified by an independent operator who did not perform the assay.

Fold-increase (post-vaccine titer/pre-vaccine titer), seroprotection (antibody titer ≥40), and seroconversion (≥4 fold increase) were calculated from the geometric mean antibody titers [29].

### 2.4 Statistical Analyses

An a priori sample size calculation indicated 68 participants would yield > 80% power at α< 0.05 with a medium effect size (f^2^ = .15) and two tested predictors. The effect size was estimated based on the work of Edwards et al. [18] and corresponds to at least a 1.5X increase in mean antibody titer due to the intervention. However, we were unable to meet this enrollment within the study period. Because of the need to provide vaccination within a certain period (autumn) and of yearly differences in the seasonal influenza vaccine, we were unable to expand the recruitment period. We have proceeded with the analyses of the existing participants, with the modified aim of understanding the potential effect size of resistance exercise on influenza antibody titers in older adults.

Participant characteristics were compared across the 3 groups by ANOVA; exercise performance measures were compared across the 2 exercise groups by *t*-tests. Data were first screened for normality and the presence of outliers graphically and homogeneity of variance was assessed by Levene’s test.

Antibody titers (geometric means) were base-2 logarithmically transformed and normality of residuals was confirmed through examination of P-P plots. Log_2_ titers were analyzed using random-intercepts restricted maximum likelihood mixed models with a variance components covariance structure. Models included time (3 time points), group (3 groups), and the interaction of group by time. Group differences within each timepoint were assessed though pairwise comparisons of estimated marginal means, adjusted for multiple comparisons by the method of Sidak. Fold-change in antibody titer was calculated for each time point relative to baseline titer (6-week titer/baseline titer; 24-week titer/baseline titer) and group-effects were assessed by Kruskal-Wallis Test. Titers against each of the 4 virus strains were considered separately.

All analyses were completed using IBM SPSS Statistics for Windows, Version 26. p< 0.05 was accepted as significant.

## 3. RESULTS

### 3.1 Participants and exercise

Twenty-nine participants (20 women) were randomized into one of three groups: No-Ex, Ex-S, and Ex-OP. Participant characteristics are shown in Table 1. Groups did not differ in these characteristics.

**Table 1.** Participant characteristics.

| **N (Female):** | **All** | **No-Ex** | **Ex-S** | **Ex-OP** | **p** |
| --- | --- | --- | --- | --- | --- |
|  | **29 (20)** | **10 (7)** | **10 (7)** | **9 (5)** |  |
| Age (years) | 73.9 ± 5.3 | 70.9 ± 5.7 | 75.8 ± 5.0 | 75.1 ± 5.3 | 0.132 |
| Height (cm) | 165 ± 6.8 | 168 ± 8.4 | 165 ± 7.0 | 164 ± 4.5 | 0.525 |
| Weight (kg) | 75.0 ± 14.6 | 75.0 ± 14.6 | 80.7 ± 16.4 | 69.7 ± 9.9 | 0.269 |
| Body mass index (kg/m<sup>2</sup>) | 27.3 ± 4.9 | 26.2 ± 4.6 | 29.7 ± 5.7 | 25.8 ± 3.7 | 0.189 |
| Grip Strength dominant (kg) | 27.4 ± 8.5 | 29.9 ± 10.5 | 27.1 ± 5.7 | 24.9 ± 8.7 | 0.450 |
| Grip Strength<br>nondominant<br>(kg) | 25.1 ± 6.9 | 28.9 ± 9.6 | 23.7 ± 3.7 | 22.8 ± 5.3 | 0.123 |

Values are mean ± S.D.; p value is associated with F tests comparing No-Ex, Ex-S, and Ex-OP

Ex-S and Ex-OP completed a total of 50 repetitions of the lateral raise and bicep curl exercises. In both groups, the weight was lowered from the 80% of 1RM used at the start of the intervention as participants fatigued and technique suffered. Ex-S and Ex-OP did not differ in exercise performance (Table 2).

**Table 2.**
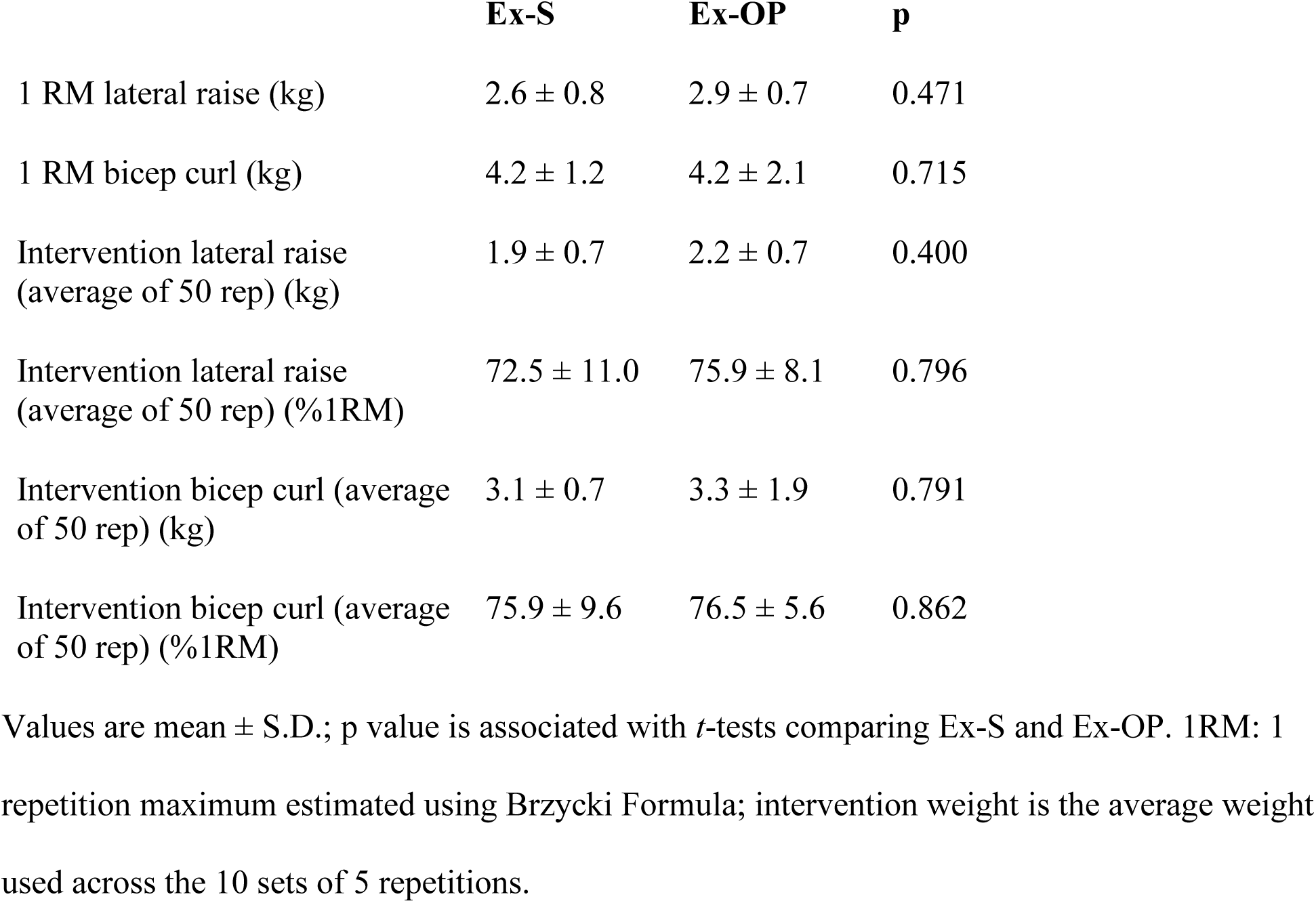
Resistance exercise performed by Ex-S (exercise in non-dominant arm) and Ex-OP (exercise in dominant arm)

|  | **Ex-S** | **Ex-OP** | **p** |
| --- | --- | --- | --- |
| 1 RM lateral raise (kg) | 2.6 ± 0.8 | 2.9 ± 0.7 | 0.471 |
| 1 RM bicep curl (kg) | 4.2 ± 1.2 | 4.2 ± 2.1 | 0.715 |
| Intervention lateral raise<br>(average of 50 rep) (kg) | 1.9 ± 0.7 | 2.2 ± 0.7 | 0.400 |
| Intervention lateral raise<br>(average of 50 rep) (%1RM) | 72.5 ± 11.0 | 75.9 ± 8.1 | 0.796 |
| Intervention bicep curl (average<br>of 50 rep) (kg) | 3.1 ± 0.7 | 3.3 ± 1.9 | 0.791 |
| Intervention bicep curl (average<br>of 50 rep) (%1RM) | 75.9 ± 9.6 | 76.5 ± 5.6 | 0.862 |

Values are mean ± S.D.; p value is associated with *t*-tests comparing Ex-S and Ex-OP. 1RM: 1 repetition maximum estimated using Brzycki Formula; intervention weight is the average weight used across the 10 sets of 5 repetitions.

Participants were asked to report the presence of any soreness in the vaccinated and unvaccinated arms for each of 7 days post-vaccination. Eight of 29 participants (3 in No-Ex, 3 in Ex-S, and 2 in Ex-OP) reported soreness in the vaccinated arm on at least one day; all reports of soreness resolved within 6 days. One participant in Ex-OP also reported soreness in the non-vaccinated arm (the arm that exercised); this soreness also resolved within 6 days.

### 3.2 Antibody titer

Serum collected at baseline, 6-weeks, and 24-weeks post-vaccination was assessed by hemagglutinin inhibition assays to determine antibody titers to the four influenza strains included in the vaccine. Geometric mean antibody titers are displayed in Figure 3. No-Ex, Ex-S, and Ex-OP did not differ in antibody titer to any strain across the three time points: A/H1N1, F(4,48.5) = 0.466, p = 0.760; A/H3N2, F(4,48.79) = 0.893, p = 0.475; B/Colorado/06/2017, F(4,48.98) = 1.091, p = 0.371; and B/Phuket/3073/2013, F(4, 48.74) = 0.307, p = 0.872.

**Figure 3.**
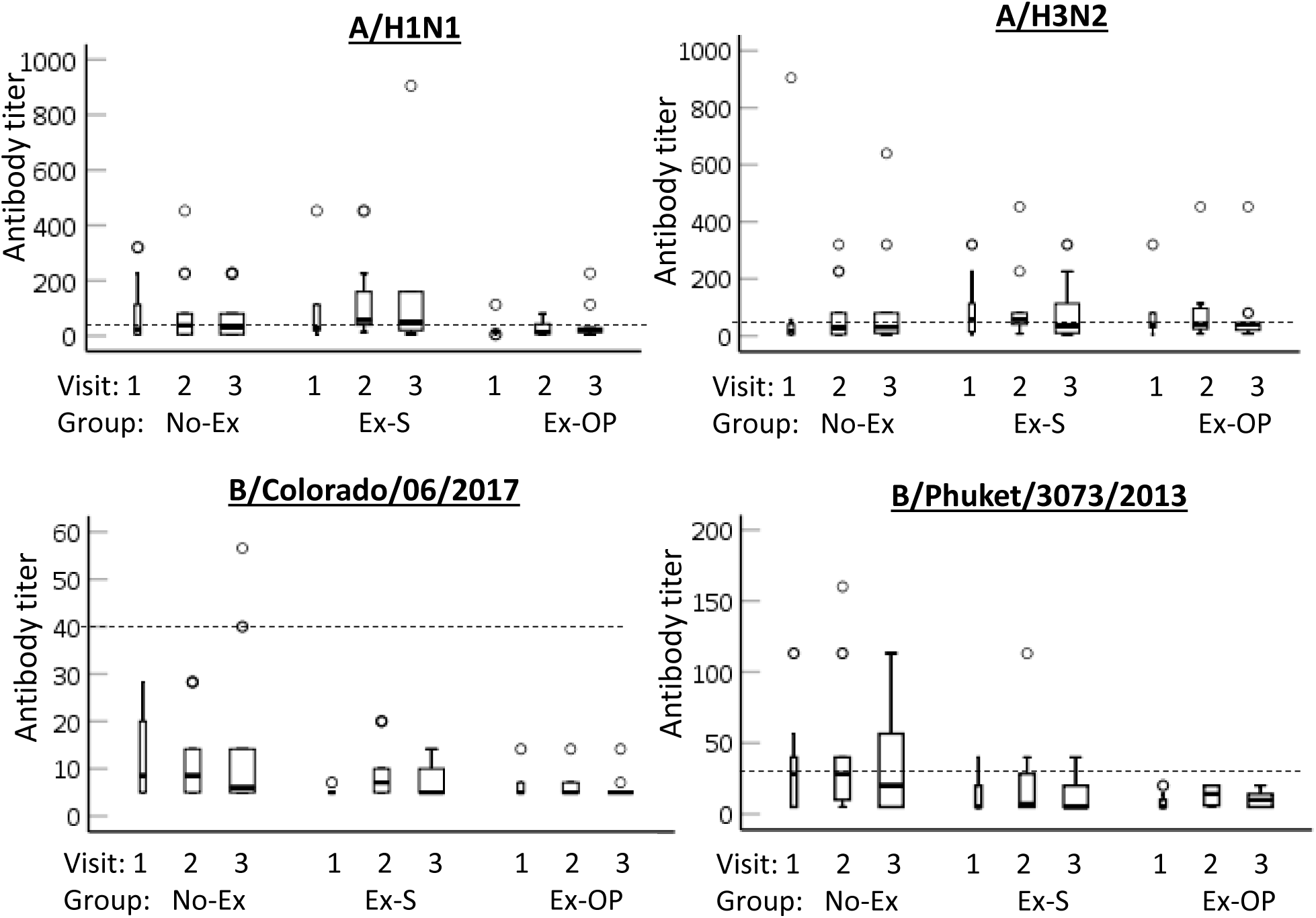
Group assignment did not impact antibody titers. Data shown are geometric mean antibody titers for each strain by visit and group. Visit 1 = baseline, Visit 2 = 6-weeks post-vaccination, Visit 3 = 24-weeks post-vaccination. Median (black bar), 1st and 3rd quartiles (box), confidence interval (error bar) and outliers (circles) are shown. Dashed line indicates seroprotection.

There was also no difference in antibody titer between the three visits, regardless of group assignment: A/H1N1, F(2,48.52) = 0.522, p = 0.597; A/H3N2, F(2,48.82) = 0.357, p = 0.701; B/Colorado/06/2017, F(2,49.01) = 1.195, p = 0.311; and B/Phuket/3073/2013, F(2, 48.76) = 2.261, p = 0.115. Accordingly, rates of seroprotection (antibody titer ≥40) remained low, particularly for B/Colorado/06/2017 (Figure 3) (Table 3).

**Table 3.** Number and proportion of all participants exhibiting seroprotection and seroconversion 6-weeks and 24-weeks post-vaccination.

|  | At 6-weeks: |  |  |  | At 24-weeks: |  |  |  |
| --- | --- | --- | --- | --- | --- | --- | --- | --- |
|  | Seroprotection |  | Seroconversion |  | Seroprotection |  | Seroconversion |  |
|  | n=26 |  | n=25 |  | n=29 |  | n=28 |  |
| Antigen | n | % | n | % | n | % | n | % |
| A/H1N1 | 14 | 53.8 | 4 | 16.0 | 13 | 44.8 | 2 | 7.1 |
| A/H3N2 | 15 | 57.7 | 3 | 12.0 | 16 | 55.1 | 3 | 10.7 |
| B/Colorado/06/2017 | 0 | 0.0 | 2 | 8.0 | 2 | 6.9 | 1 | 3.6 |
| B/Phuket/3073/2013 | 6 | 23.0 | 4 | 16.0 | 5 | 17.2 | 2 | 7.1 |

Seroprotection: antibody titer $\geq 40$ ; seroconversion: fold-change from pre-vaccination $\geq 4$ .

To understand if groups differed in the change in antibody titer from baseline, we compared fold-change at 6-weeks and 24-weeks between groups (Figure 4). Overall, rates of seroconversion (fold-change ≥4) were low (Table 3). No group differences in 6-week or 24-week fold change were noted for any virus strain (Table 4). However, a large effect size was noted for fold-change at 6-weeks for B/Colorado/06/2017. Visual inspection of the data suggests a trend for a greater fold-change in Ex-S (Figure 4). Mean fold increases at 6-weeks in response to B/Colorado/06/2017 were 1.08 (No-Ex), 1.93 (Ex-S), and 1.06 (Ex-Op).

**Figure 4.**
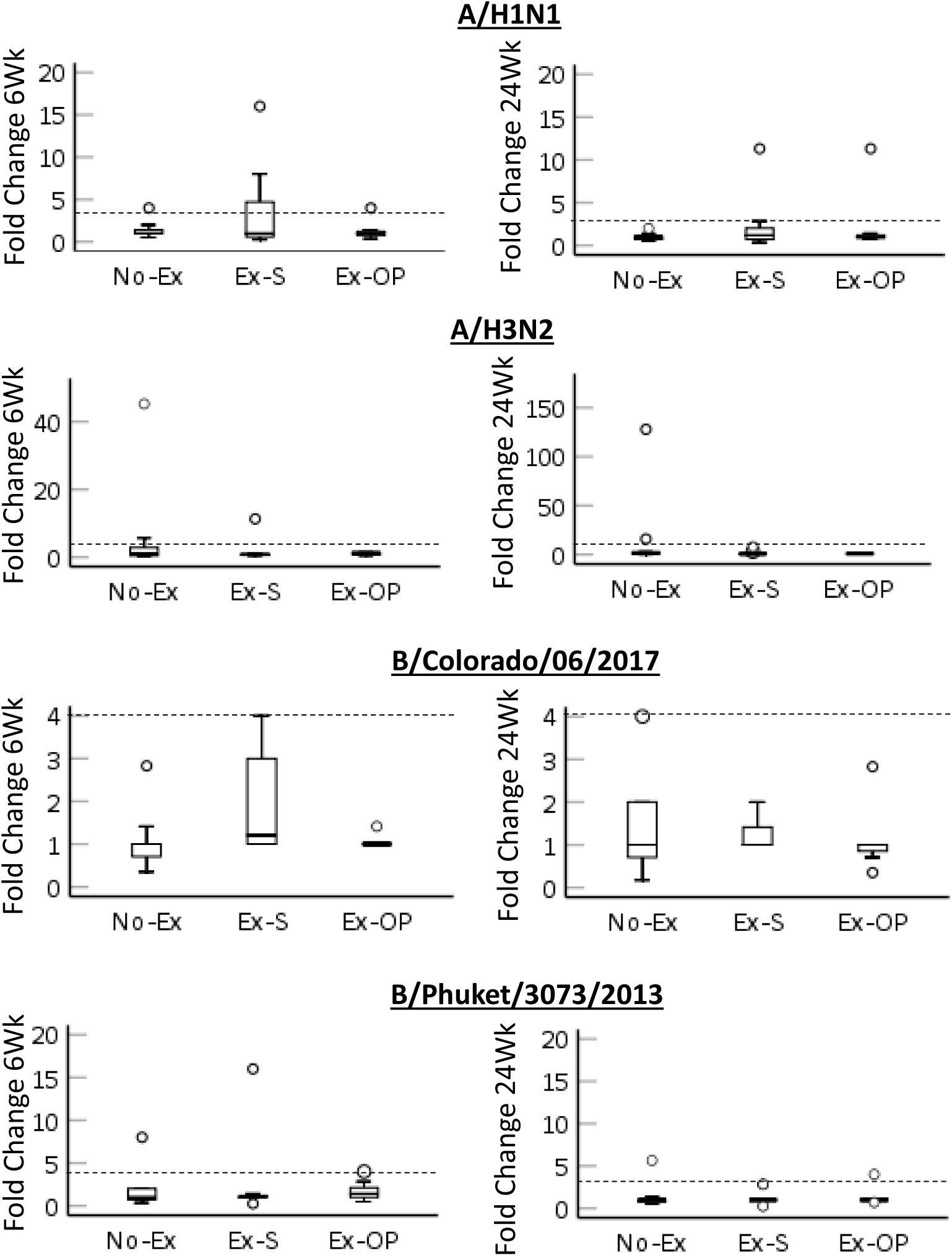
Fold change in antibody titer from baseline 6-weeks post-vaccination (left) and 24-weeks post-vaccination (right) for each strain by group. Median (black bar), 1st and 3rd quartiles (box),confidence interval (error bar) and outliers (circles) are shown. Dashed line indicates seroconversion.

**Table 4.**
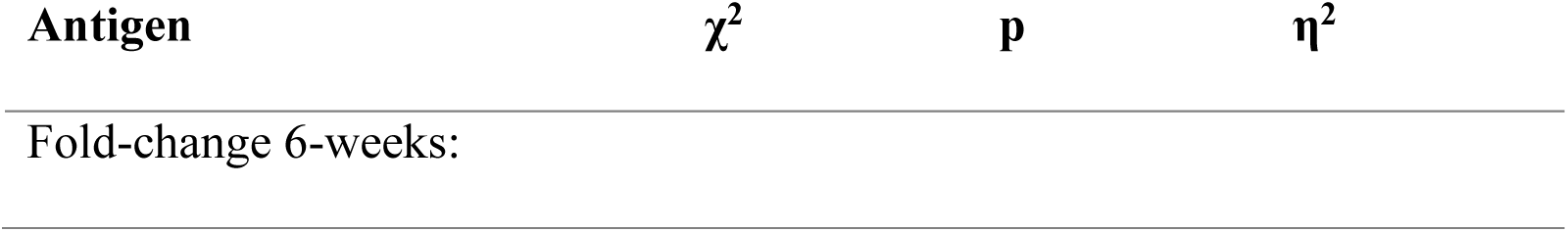
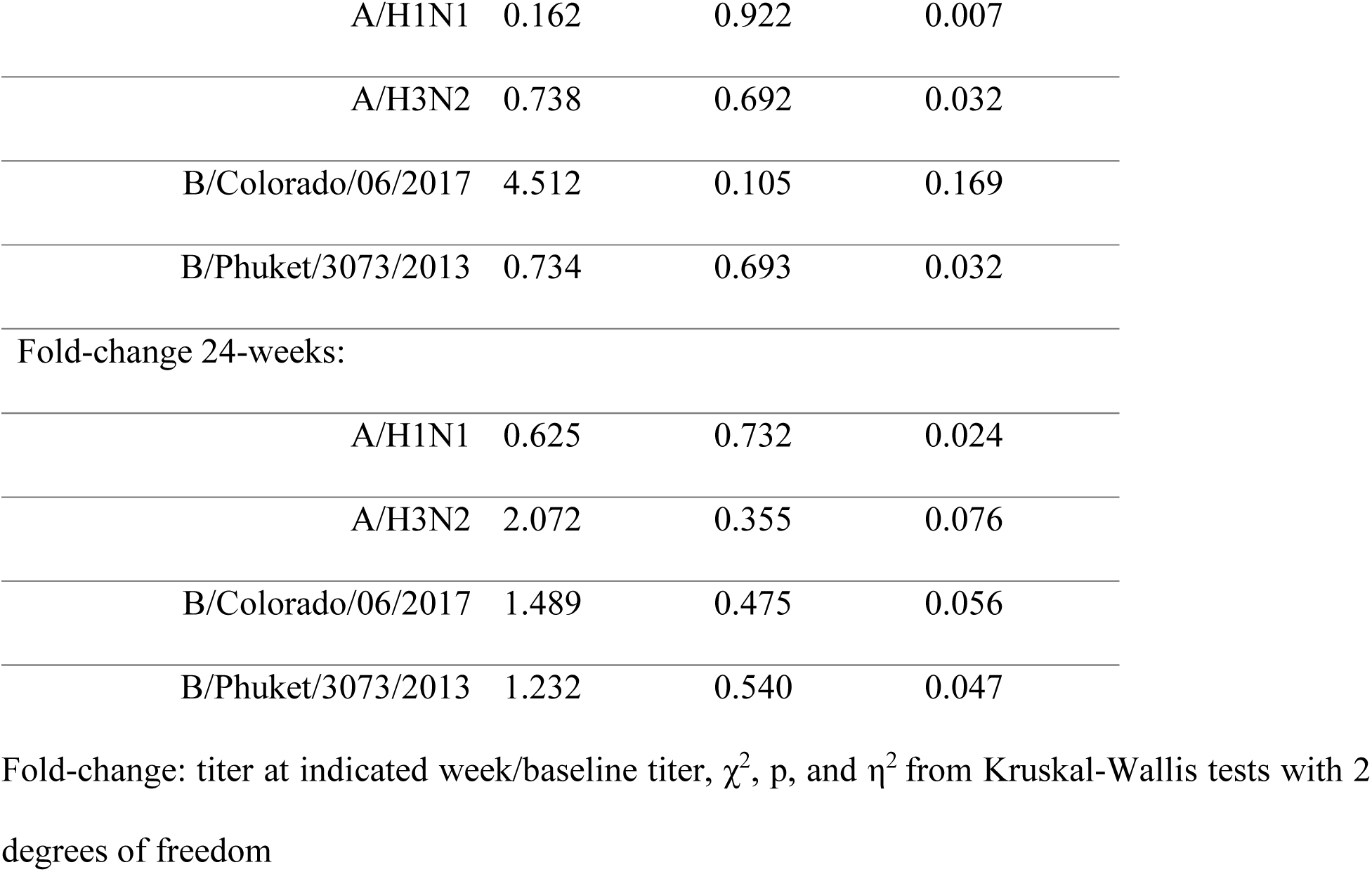
Effect of group in fold-change in antibody titers 6-weeks and 24-week post-vaccination.

Despite the low rates of seroprotection, only one participant (Ex-S) reported flu-like symptoms 18 weeks post-vaccination; influenza infection was not clinically confirmed in this participant.

## 4. DISCUSSION

To the best of our knowledge, this study is the first to examine the effect of a single session of eccentric-focused resistance exercise on influenza vaccine responses in older adults. Older adults frequently have insufficient responses to the influenza vaccine and so are at increased risk for influenza infection. We hypothesized that eccentric exercise performed in the same arm subsequently vaccinated would improve protective immune responses to the vaccine. Our primary outcome of interest, hemagglutinin inhibition antibody titers, was compared between three groups before and after (6-weeks and 24-weeks) vaccination with the seasonal quadrivalent influenza vaccine. Contrary to our hypothesis, no differences were found between No-Ex, Ex-S, and Ex-OP across this timeframe, indicating that an acute bout of unfamiliar eccentric resistance exercise, performed just before vaccination either in the same or opposite arm, did not influence antibody responses to the vaccine in this population of older adults.

Observations that greater physical activity levels are associated with greater immune responses following influenza vaccination have provided a rationale for chronic exercise training interventions as a means to improve vaccine responses [30–34]. Although 5–10 month long exercise training interventions have had moderate success in enhancing anti-influenza antibody titers [35–37], the time commitment required may render such interventions impractical for broad implementation. Single sessions of exercise have also been proposed as a means of enhancing immune responses to vaccination. A moderate intensity aerobic exercise bout performed prior to inoculation yielded higher influenza antibody titers in young women, but not young men, 4-weeks and 20-weeks post-vaccination [38]. In a subsequent examination by the same group it was reported that eccentric resistance exercise prior to influenza vaccination also significantly increased antibody titers 6-weeks post-vaccination by approximately 1.5X the control condition in young women, corresponding to a medium effect size [18]. Though the current study mirrored the eccentric-focused exercise protocol in the Edwards et al. study, effect sizes reported in the current study are small in almost all cases. Key differences which may explain this discrepancy in results include the fact that the study by Edwards et al delayed vaccination for 6 hours after the exercise bout, whereas the current study vaccinated participants immediately (< 5 minutes) post-exercise. Delaying vaccination for several hours after exercise may allow a greater immune response to the exercise bout to develop, although local and systemic immune changes are also noted immediately

[16]. A study aiming to optimize the timing of the bout of eccentric exercise directly compared vaccinating immediately after exercise or delaying for 6 hours, but found no difference between any groups in the intervention, including the no-exercise control [39].

A second difference is that the study by Edwards et al used an exercise intensity of 85% of 1RM for the intervention, which was maintained for all 50 repetitions. The participants in the current study instead were only able to maintain an average intensity of ∼70–75% 1RM across the 50 repetitions. We selected a slightly lower goal intensity at the start of the intervention (80% 1RM) in consideration of joint health of the older, resistance-trained naïve participants and lowered intensity as technique suffered. An alternative strategy could have been to maintain weight but increase intra- and inter-set rest. Self-reports of muscle soreness were low following our intervention and did not differ in frequency from the No-Ex group, suggesting both that the exercise protocol was well tolerated in this population and that our exercise protocol may not have stimulated a local inflammatory reaction. The lack of a systemic marker of muscle damage (e.g. creatine kinase) is a limitation to our study. However, these differences in intensity may not have influenced antibody titers, as was demonstrated in a direct comparison of the effect of eccentric arm exercise at 60%, 85%, and 110% of 1RM on antibody responses to influenza vaccination that found no difference between exercise groups [20].

Perhaps the largest difference between the earlier eccentric-exercise investigations and the current study is the fact that our participants were older adults, rather than younger adults who typically achieved clinically protective immune responses to the vaccine even when assigned to the resting control group [18–20,39]. In contrast, vaccination of the older participants here largely failed to generate seroconversion, similar to other reports [5,8]. We had hypothesized that exercise would provide a greater benefit in the current study, as others have found that exercise can enhance immune responses, especially in cases where the control response is weak [18–20,39]. Although the exercise effect is not statistically significant, our data offer some support of this idea. There was a trend for an effect of group on fold-change in antibody titer at 6-weeks for B/Colorado/06/2017, apparently due to the larger increase in antibody titer in Ex-S. The B/Colorado/06/2017 strain appeared to be the least immunogenic, as the antibody titers generated to this strain were the lowest on average of the four strains in the vaccine.

The mechanisms underlying exercise-induced improvements in vaccine responses are unknown but may act through the recruitment of immune cells, particularly antigen presenting cells, into local muscle. Eccentric exercise, particularly in those not accustomed to the exercise, results in neutrophil and macrophage infiltration into the exercised muscle which release reactive oxygen and nitrogen species and cytokines [16,40–42]. This is similar to the response to chemical adjuvants added to vaccines to enhance immune responses, as like eccentric exercise, chemical adjuvants initiate strong inflammatory reactions and immune cell recruitment at the site of the injection[43,44]. Thus, eccentric resistance exercise may act as an adjuvant through localized inflammation at the site of the vaccine injection. Improvements in antibody-responses to vaccination after cardiorespiratory exercise also suggest that systemic effects of exercise may play a role in immune enhancement [34,38]. Exercise is well known to elicit a transient increase in leukocytes and cytokines in circulation [45]. As resistance exercise protocols used to date have only included exercise of the arm to be inoculated, it is impossible to separate the local and systemic exercise effects. Comparing cardiorespiratory and resistance exercise protocols is also unsatisfactory, due to difficulties in matching exercise intensity. Although we aimed to overcome these limitations in the current study by including a group matched in exercise but performed in the dominant (and therefore unvaccinated) arm (Ex-OP), the small sample prevents meaningful comparison. Importantly, both local and systemic immune responses to exercise are reduced in older adults [46]. Intensity-matched cardiorespiratory exercise yields a smaller increase in leukocytosis in older adults relative to young adults [46,47]. Although age-related declines in skeletal muscle mass and strength may leave older adults more vulnerable to exercise-induced muscle damage [48], eccentric exercise-induced increases in circulating and muscle-infiltrating neutrophils and plasma IL-6 are reduced in older adults [49–51]. Thus, it may be that the older adults in the current study did not generate as strong of an immune response to the exercise as younger participants in earlier studies using a similar exercise protocol.

A major limitation of the current study is the small number of participants relative to the actual effect size, which decreased power. The primary reason provided by interested individuals who declined participation was the convenience in receiving their seasonal influenza vaccine from their usual health care provider compared to the time required for participation. This suggests individuals are not opposed to resistance exercise *per se* prior to vaccination, and perhaps recruitment would be higher if it could be performed in the clinic waiting room. An additional reason for screening exclusion was for participation in the prior 6-months in arm resistance exercises. Participants were required to be arm resistance-training naïve to avoid heterogeneity in response due to the repeated-bout effect [16,17] but this did remove a segment of the older adult population that may be most receptive to performing arm resistance exercise prior to vaccination. Further studies are needed to understand if resistance-trained individuals respond differently from resistance-naïve individuals in the effects on vaccination.

In conclusion, we report that an acute bout of unaccustomed eccentric exercise performed before vaccination with the 2018–2019 quadrivalent influenza vaccine did not yield differences in geometric mean titer, fold-increase, seroconversion, or seroprotection in older men and women, relative to a resting, standard-of-care control. Although the participants were naïve to the exercise, it was well-tolerated. The current study is limited by the small number of participants. Thus, we view these results as providing guidance for future studies targeting the use of exercise as a means of enhancing immune responses to influenza vaccine in older adults.

## Data Availability

Data referred to in the manuscript are available upon request.

## ACKNOWLEDGEMENTS

This research was conducted with the support of the National Institute on Aging (Grant R03AG052778, E.L. Principal Investigator, R.S., M.M., and M.L., Co-Investigators).

## AUTHOR CONTRIBUTIONS

EL, RJ, and MM designed the study; EL, MT, and MM supervised the study; EL, MT, and ML performed data analyses. All authors contributed to manuscript writing.

